# Mpilo Outcomes for Stroke Study (MOSS): Does taking antihypertensives prior to stroke affect mortality during acute stroke admissions?

**DOI:** 10.1101/2021.07.01.21259883

**Authors:** Nigel Gwini, Desmond Mwembe, Rudo Gwini

**Author notes:** **Corresponding Author:** Nigel Gwini, MD (NewYork-Presbyterian Medical Center, NY USA). **Co-author:** Mr. Desmond Mwembe (NUST Faculty of Statistics, Bulawayo, Zimbabwe). **Principal Investigator:** Dr. Rudo Gwini (Mpilo Central Hospital, Bulawayo, Zimbabwe and NUST Faculty of Medicine).

## Abstract

**Objective:** To assess if taking outpatient antihypertensives prior to stroke onset influences acute stroke inpatient mortality.

**Design:** Retrospective chart review of 417 adults admitted for acute stroke from January 2013 to December 2014.

**Setting:** Single tertiary hospital in Zimbabwe.

**Participants:** Of the 417 adult patients admitted with stroke, 40 were excluded (2 rheumatic heart disease, 10 transient ischemic attacks, and 28 incomplete documentation).

**Methods:** Study registry was obtained from acute stroke billing codes in the hospital database and SPSS 23 was used for analysis.

**Primary and secondary outcomes and measures:** Inpatient mortality was the primary outcome and determination of reported outpatient antihypertensive regimens was the secondary outcome.

**Results:** Among the 377 patients in the final analysis the mean age was 65.8 ± 15.7 years, 64.5% were female, 68.9% had known hypertension, and 51.2% were taking outpatient antihypertensives. Overall inpatient mortality was 134 (35.5%, 95 CI: 30.6 -39.6). Mortality was similar among patients taking and not taking outpatient antihypertensives, 39.2% and 31.5% respectively (*X*^*2*^ *test, p=0*.*4;* 95 CI 31.3-40.3). Four most common antihypertensive regimens were: calcium channel blocker/thiazides dual therapy, thiazide monotherapy, calcium channel blocker monotherapy, and calcium channel blocker/ angiotensin converting enzyme inhibitor or angiotensin receptor blocker dual therapy.

**Conclusions and Relevance:** Only half of the patients admitted for stroke were taking antihypertensives prior to stroke onset. There was no statistical significance in inpatient mortality associated with taking antihypertensives prior to developing stroke, suggesting either the limited impact of outpatient antihypertensives on in-hospital mortality or low outpatient antihypertensive adherence.

**Strengths and Limitations of the Study:**

- We studied a large group of patients at a major safety net center in a region of Zimbabwe that is chronically under-represented in stroke studies
- The degree of outpatient hypertension control among patients on antihypertensives could not be determined due to lack of access to outpatient records
- Poor medical knowledge, altered mental status from stroke, and the absence of an integrated outpatient pharmacy database limited accurate medication reconciliations
- Lack of neuroimaging data limited the ability to discern between ischemic versus hemorrhagic strokes and rule out structural stroke mimics such as intracranial tumors
- Inconsistent and incomplete documentation practices such as omission of vital sign documentation on admission limited data collection

**Funding Statement:** This research received no specific grant from any funding agency in the public, commercial or not-for-profit sectors.

**Competing Interests:** None of the authors have any competing interests to declare.

**Statement of Ethics Approval:** This research was approved by the ethics body of Mpilo Central Hospital on 21 June 2016 as part of the mandate to assess The Burden of Hypertension and its Co-morbidities in Patients Admitted at Mpilo Central Hospital.

## Introduction

Stroke is one of the leading causes of death and disability according to the World Health Organization (WHO).^***1***^ Cerebrovascular accidents disproportionately impact low-earning and impoverished communities. According to the 2010 Global Burden of Disease (GBD) study, three quarters of patients diagnosed with initial strokes resided in low-to middle-income countries (LMICS) such as Zimbabwe.^***2***^ Furthermore, almost ninety percent of all stroke-related mortality occurred in LMICS.^***3***-***4***^ The high rates of disability and death secondary to cerebrovascular accidents have established stroke as a major global public health issue.

### Definition, diagnosis, and management of acute stroke

World Health Organization (WHO) defines, a cerebrovascular accident or stroke as a state of “rapidly developing clinical signs of focal (or global) disturbance of cerebral function, lasting more than 24 hours or leading to death, with no apparent cause other than that of vascular origin.” ^***5***^ Diagnosis of an acute stroke is two-fold. First, it involves a thorough physical examination with accompanying neuro-imaging (CT/MRI) if available. Second, a parallel workup to exclude stroke mimics e.g. hypoglycemia or infection is conducted to rule out non-vascular etiologies of focal neurological deficits.^***6***^ In high-resource settings, CT head imaging is used to exclude acute intracranial hemorrhage or structural abnormalities that would preclude safe delivery of tissue plasminogen activator (tPA) for an acute ischemic stroke.^***7***^

### Prior Stroke Research in Sub-Saharan Africa

Prior stroke studies in Sub-Saharan Africa, specifically in Zimbabwe, have focused on the following: stroke risk factor identification, stroke incidence, and inpatient quality metrics such as length of hospitalization and inpatient mortality.^***8***-***9***^ The INTERSTROKE and SIREN studies both identified hypertension as the chief modifiable risk factor for developing stroke.^***4***,***10***^ Hypertension leading to strokes in Zimbabweans has been documented since the 1980s.^***11***^ Most recently, a multi-center investigation at three teaching hospitals in Harare showed that approximately 60% of stroke admissions had known hypertension.^***8***^ In this cohort, overall in-hospital mortality from stroke was 25%.

To our knowledge, there are no Sub-Saharan studies in the literature examining the differences in acute stroke mortality outcomes for patients on antihypertensive medications prior to acute stroke compared to those not taking medications. The landmark North American Antihypertensive and Lipid-Lowering Treatment to Prevent Heart Attack Trial (ALLHAT) study, a prospective randomized control trial, assessed the efficacy of multiple antihypertensive monotherapy regimens in preventing major adverse cardiac events such as stroke.^***12***^ The ALLHAT study also examined all-cause mortality during the study, but it was not designed to capture the data for inpatient mortality from acute stroke nor did it account for patients on as many as three or more antihypertensive medications.

This retrospective chart review was designed to determine if there were any differences in inpatient mortality from acute stroke among subjects taking or not taking outpatient antihypertensive medications prior to stroke onset. Furthermore, this study intended to characterize the specific antihypertensive regimens that subjects were taking prior to acute stroke onset.

## Methods

### Study Design

This study was a retrospective chart review conducted at a tertiary referral 1,000-bed center in Zimbabwe. An electronic search using ICD-10 code I63 (cerebral infarction) or the terms “stroke”, “cerebrovascular accident”, or “cerebral infarction” in the hospital registry was performed. A search filter for patients ages 18 and above was implemented. A total of 417 medical record numbers for adult patients ages 18 and older who were admitted for acute stroke from January 2013 through December 2014 were identified.

Each paper medical record was extracted manually from the hospital records department and reviewed by one of three clinical investigators. To meet criteria for inclusion, each patient required documented symptoms consistent with the aforementioned WHO clinical definition of stroke.^***5***^ Exclusion criteria were any alternative non-vascular explanation for focal neurological deficits.

Of the 417 patients, 40 were excluded for the following reasons: 10 subjects were determined to have transient ischemic attacks; 2 subjects had rheumatic heart disease with a high suspicion for “hypertension-independent” cardioembolic stroke from left atrial appendage thrombi; and 28 subjects had incompletely documented charts to meet full inclusion criteria. Records missing either the patient’s age or vital signs on admission were considered incomplete and excluded from the final analysis.

An assessment of the antihypertensive regimen for each of the 377 subjects in the final analysis was conducted. Each subject was assigned to one of the following five groups based on the number of antihypertensives drugs they were taking: none, one medication, two medications, three or more medications, and an unknown number of medications for patients taking antihypertensives but unable to recall their regimen (**Figure 1**).

**Figure 1.**
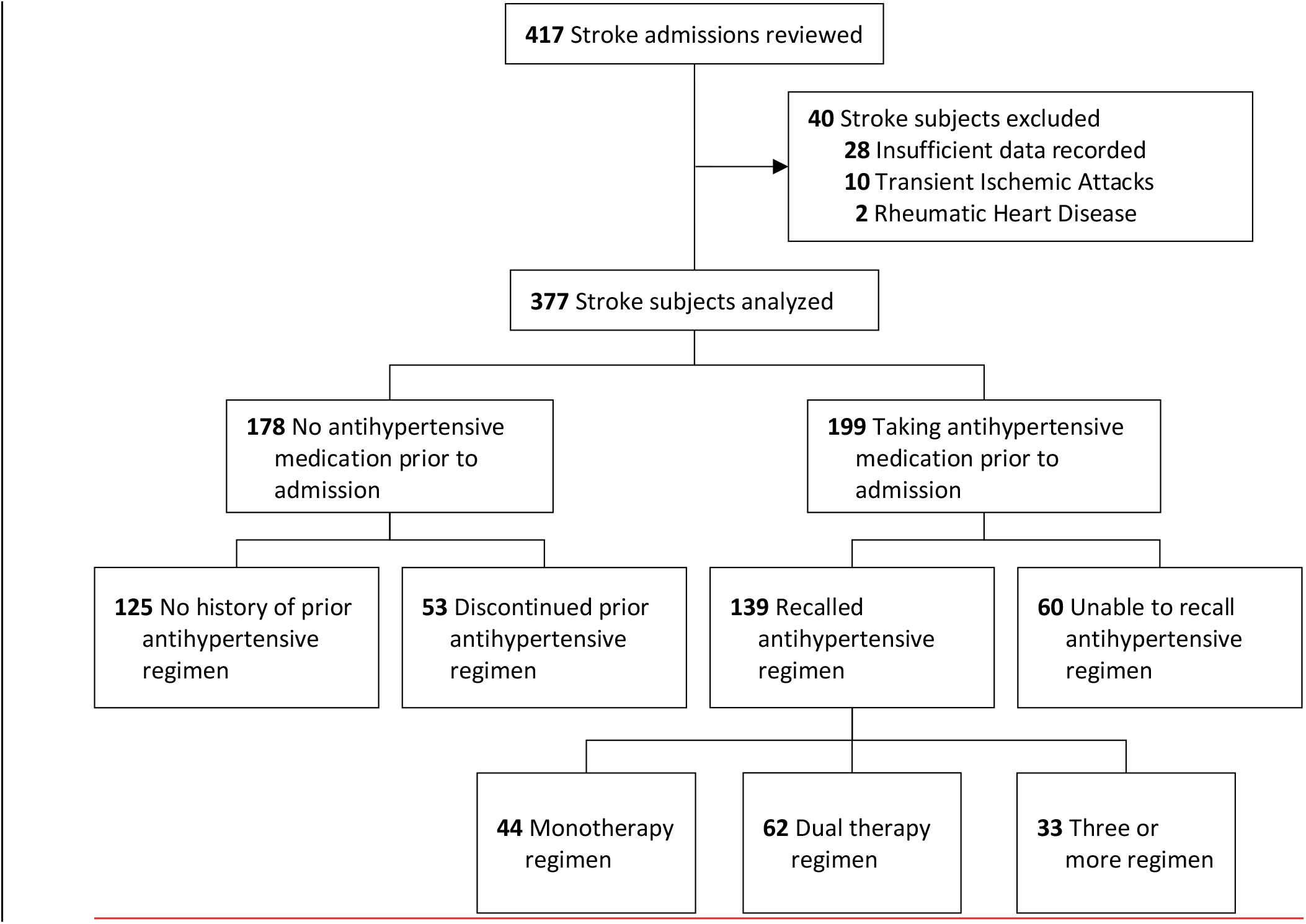
Selection of eligible subjects and assignment based on number of medications.

### Statistical Analysis

Data analysis was performed using SPSS 23 software. Mortality outcomes were calculated as percentages for the following categories: overall, use of medication, non-use of medication. 95% CI were also calculated for the proportions. Comparisons of Categorical variables between the outcomes were analyzed using *χ*^***2***^ test. An alpha value of 0.05 was chosen to achieve a 95% confidence level and a p-value <0.05 was considered statistically significant.

### Patient Disposition and Baseline Characteristics

Baseline characteristics for the 377 patients in the final analysis were summarized in Table 1. The mean patient age was 65.7 ± 16.0 (95 CI: 64.3 -67.2) and two-thirds of patients were female 243 (64.4%) (**Table 1**). A documented history of hypertension was observed in 260 (68.9%) eligible patients. A similar prevalence of hypertension across sexes: 92 (68.7%) among males and 168 (69.1%) among females was noted.

**Table 1.**
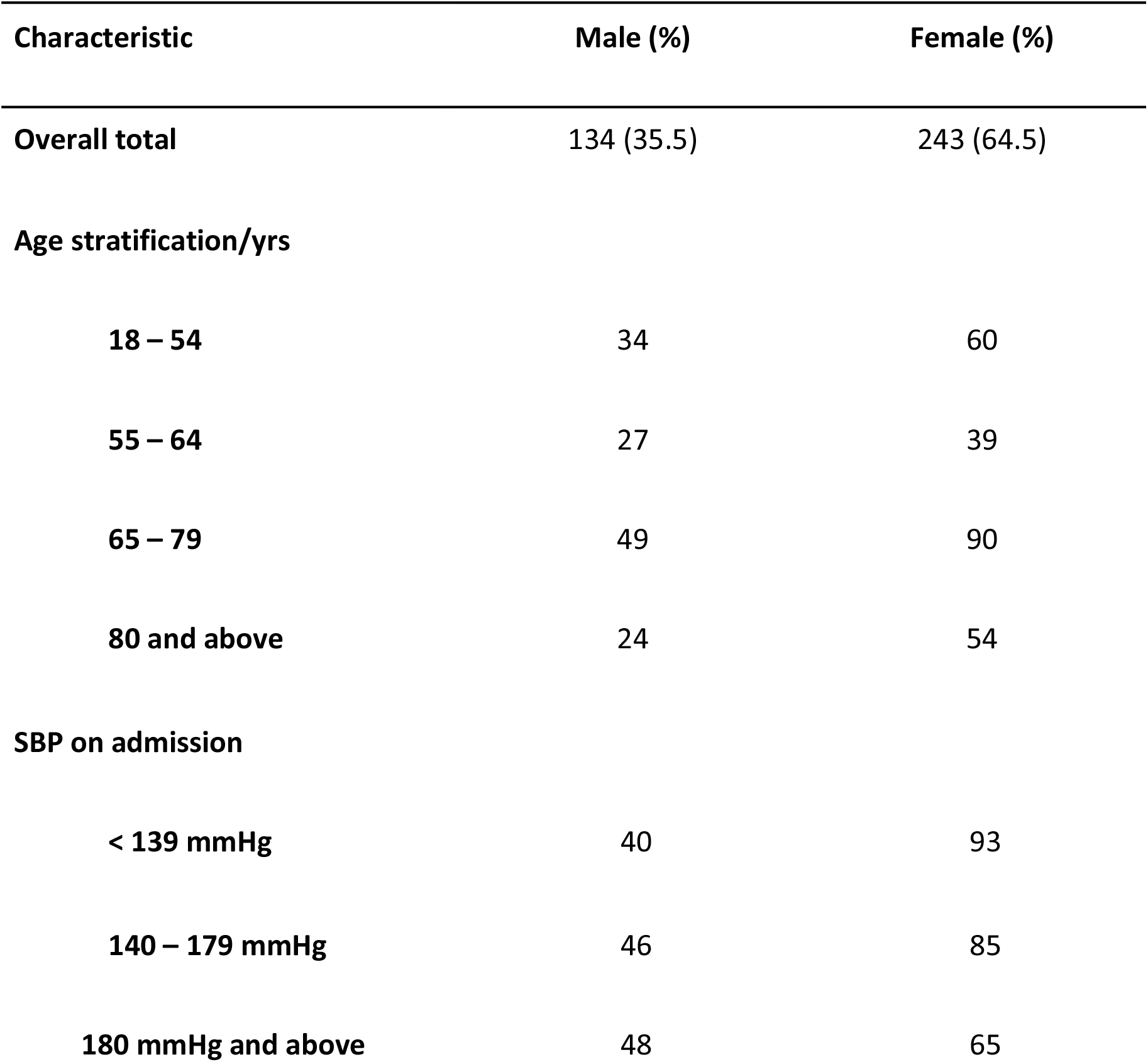
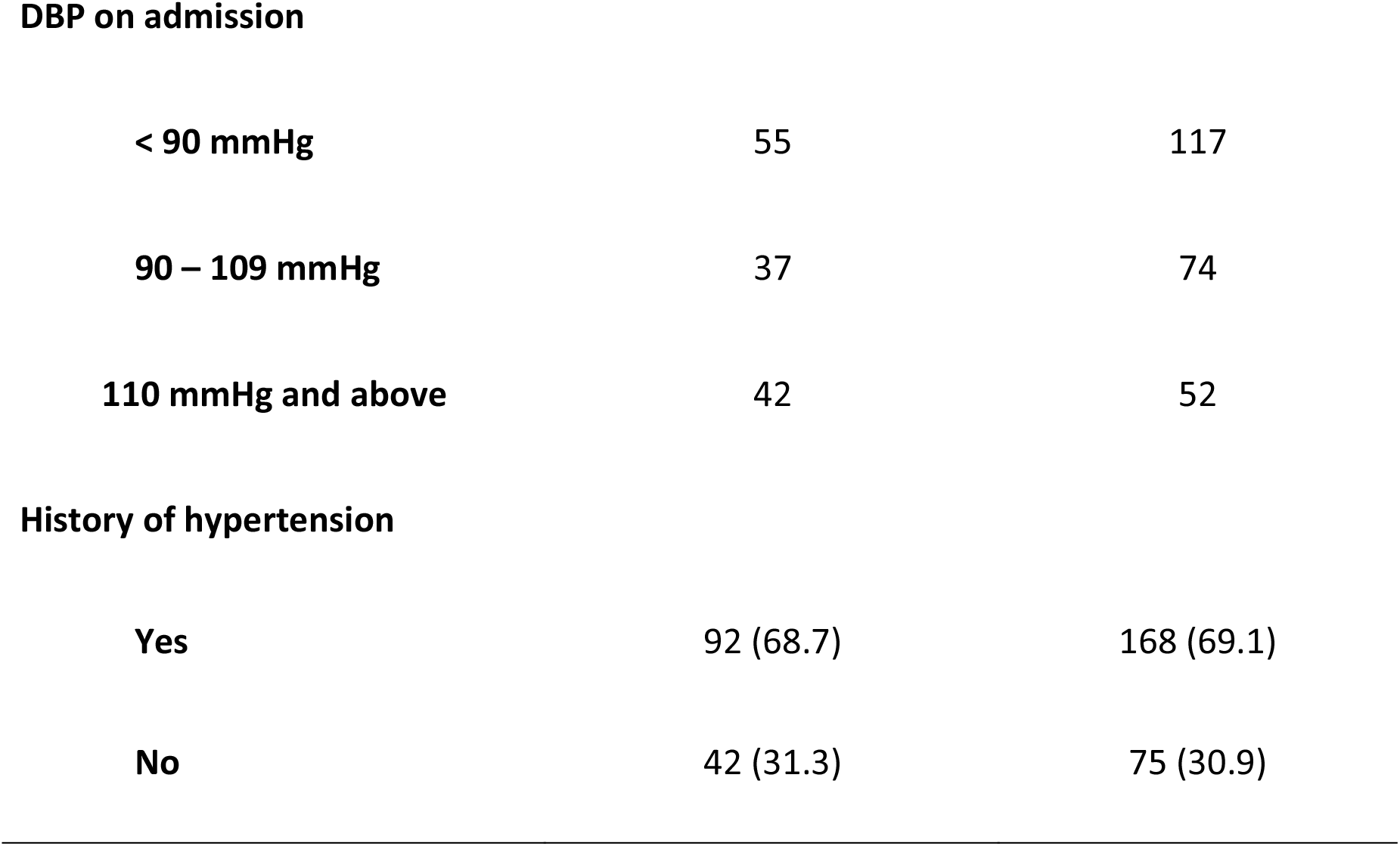
Baseline characteristics of subjects in final analysis.

Of the 377 subjects analyzed, 178 (47.2%) were not taking any antihypertensives prior to stroke onset. Specifically, 125 patients had never been on any antihypertensive medications while 53 patients had discontinued their prior antihypertensive regimen.

### Primary outcome

During the defined study period, a total of 134 (35.5%) patients died during their acute stroke hospitalization. Of the deceased, 56 (31.4%) were not on antihypertensives compared to 78 (39.2%) who were taking antihypertensives prior to admission (**Table 2**). There was no statistical difference in inpatient mortality from acute stroke among patients taking or not taking antihypertensives prior to admission (*X*^*2*^ *test, p=0*.*4;* 95 CI 31.3-40.3).

**Table 2.**
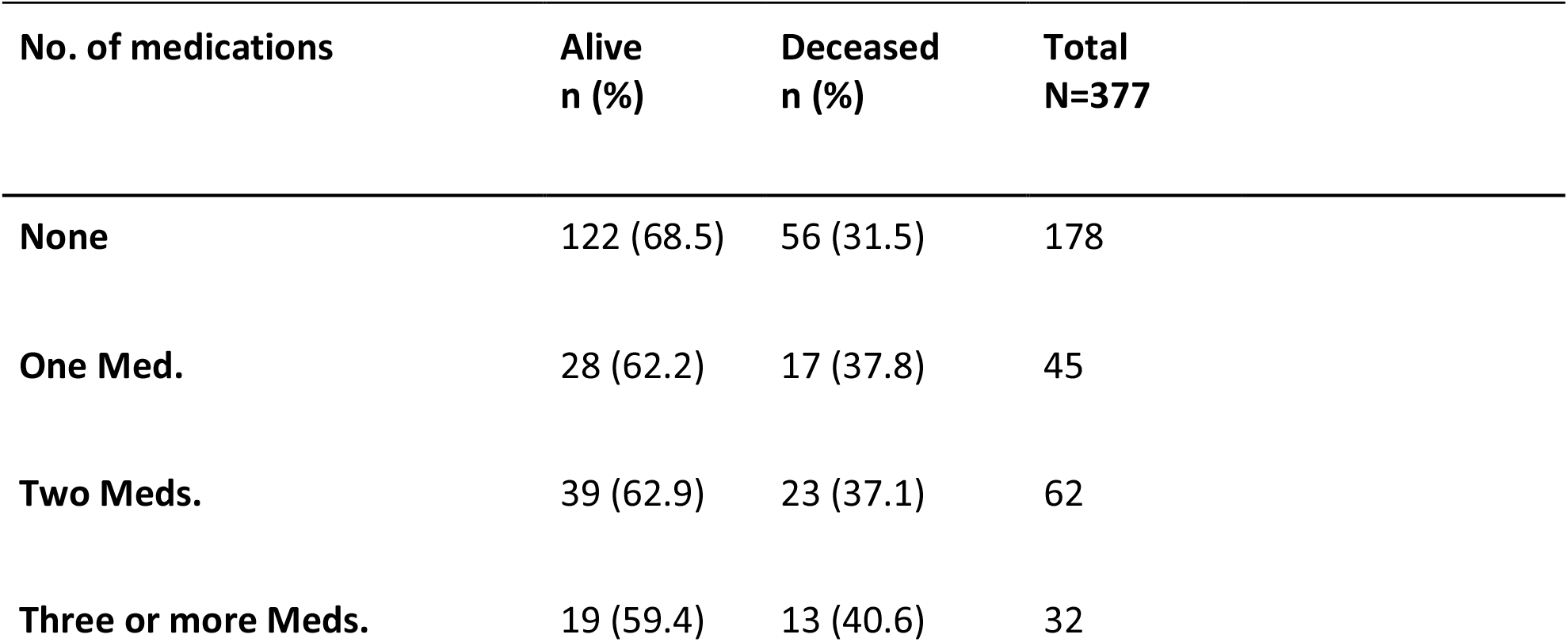
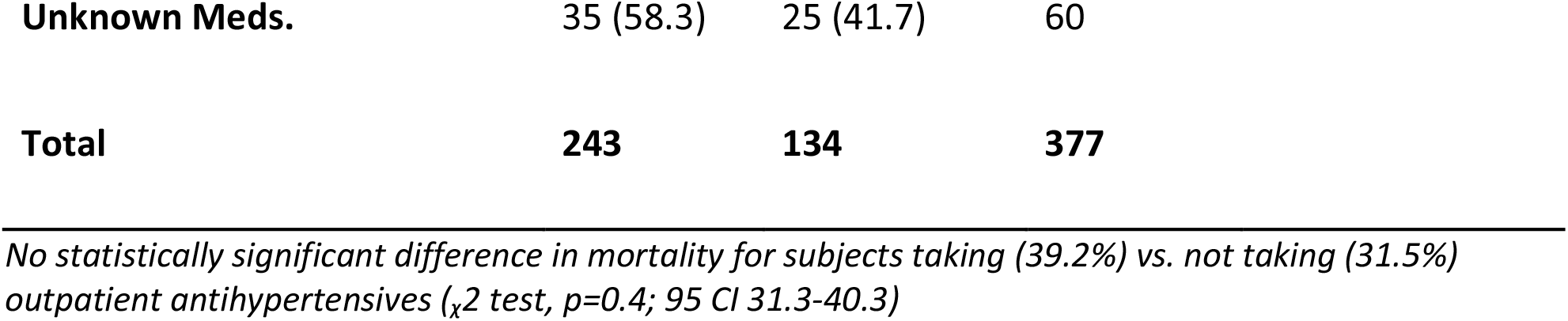
Mortality Outcomes Number of Medications.

### Mortality Outcomes by Sex and Number of Medications

Percent mortality was similar between male and female patients not taking medications, those taking unknown medications and patients taking one, two, and four antihypertensives prior to acute stroke (*respective p-values were: 0*.*126, 0*.*366, 0*.*523 and 0*.*134*). However, there was a higher mortality of 67% (n=9) among males taking three or more medications compared to 40% mortality (n=10) in females in a similar category (*p=0*.*0223*) summarized in **Table 3**.

**Table 3.** Mortality Outcomes by Sex and Number of Medications.

|  | Males | Females |  |
| --- | --- | --- | --- |
| No. of medications | Deceased<br>n (%) | Deceased<br>n (%) | ($\chi^2$ p-value) |
| None | 26 (38.2) | 30 (27.3) | 0.126 |
| One Med. | 5 (50.0) | 12 (34.3) | 0.366 |
| Two Meds. | 7 (31.8) | 16 (40.0) | 0.523 |
| Three or more Meds. | 6 (75.0) | 7 (29.2) | 0.0223 |
| Unknown Meds. | 8 (30.2) | 17 (50.0) | 0.134 |

### Mortality Outcomes by Blood Pressure on Admission

Mortality data was similar across the three systolic blood pressure cutoffs (Range: 33.6 – 36.8 % mortality) and the three diastolic blood pressure cutoffs on admission (Range: 30.9 – 37.8% mortality). There was no statistically significant increase in mortality across the admission systolic blood pressure groups (*X*^*2*^ *test, p=0*.*8*) **(Table 4)**.

**Table 4.**
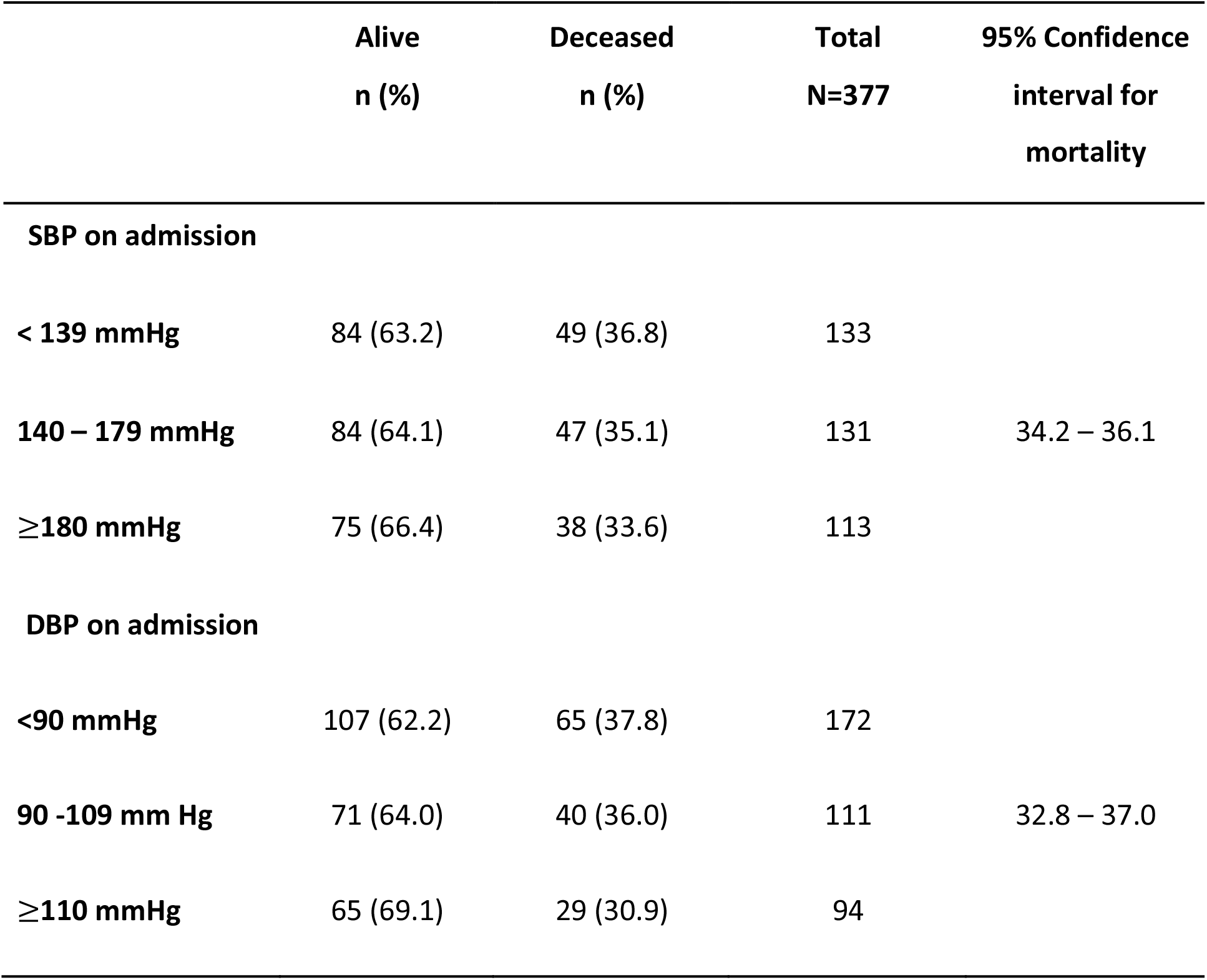
Mortality Outcomes by Blood Pressure on Admission.

### Secondary outcome: Antihypertensive regimens prior to acute stroke

Among the 199 patients taking antihypertensive medications, 44 (22.1%) were on monotherapy, 62 (31.2%) were on dual therapy, 33 (16.6%) were taking three or more antihypertensive medications, and 60 (30.2%) were unable to recall their antihypertensive regimen. Twenty-eight different combinations of medications were identified. The four most common antihypertensive regimens were: calcium channel blockers/thiazides dual therapy (31), thiazide monotherapy (28), calcium channel blockers monotherapy (13), calcium channel blockers/ angiotensin converting enzyme inhibitor or angiotensin receptor blocker dual therapy (12). All the medication combinations are summarized in **Figure 2**.

**Figure 2.**
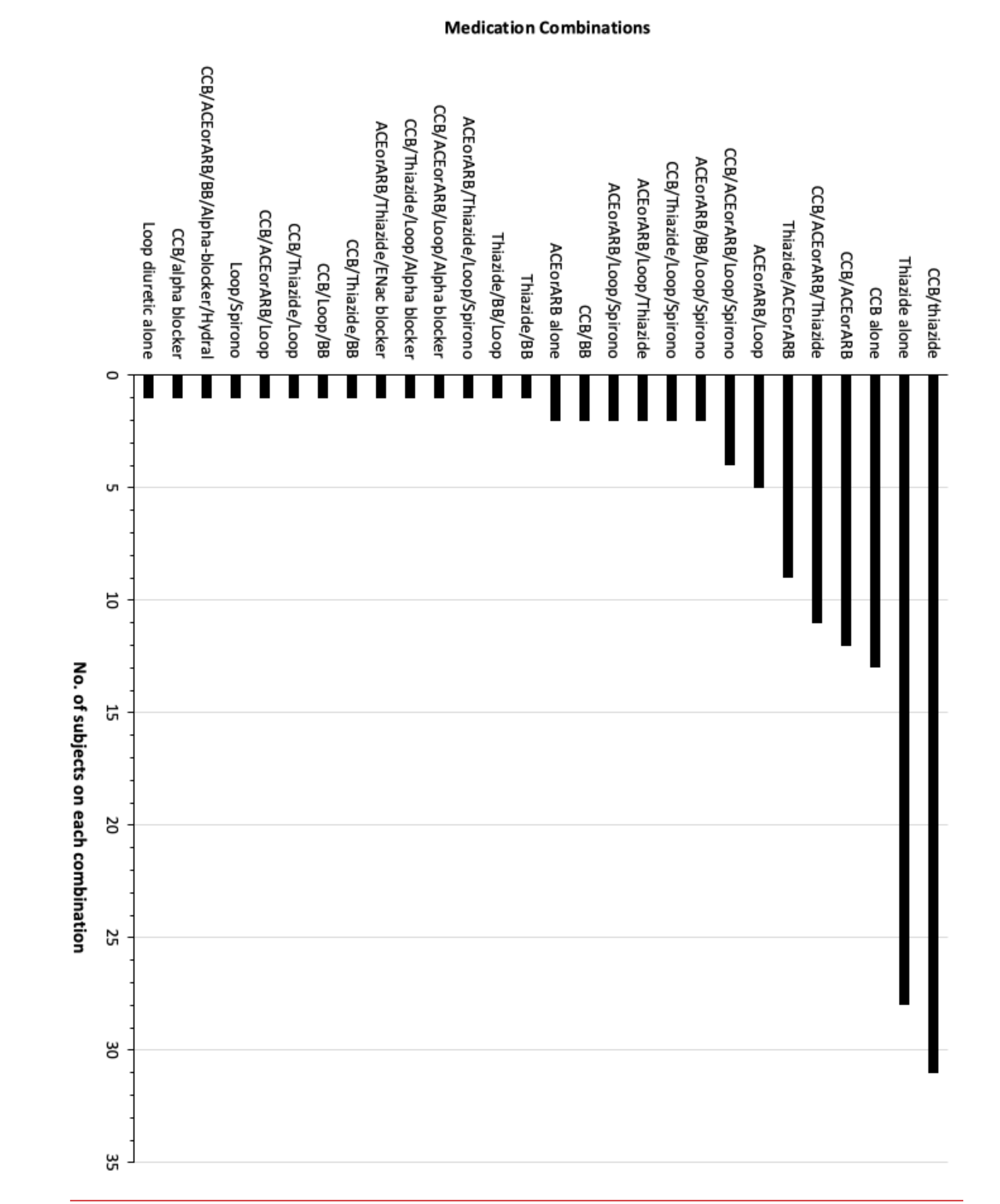
Bar Chart with Number of Patients on Each Medication Combination.

## Discussion

In this single center retrospective chart review of patients admitted for acute stroke, there was no difference in inpatient mortality observed among patients taking or not taking outpatient antihypertensives prior to acute stroke onset. Additionally, there were mostly no differences in mortality outcomes from stroke observed across sexes and admission blood pressure readings. The exception were males taking three or more outpatient antihypertensives who had a higher inpatient mortality compared to the females in a similar category but interpretation of this discrepancy is limited by the small sample sizes of these cohorts. The four most common outpatient antihypertensive regimens were: calcium channel blocker/thiazides dual therapy, thiazide monotherapy, calcium channel blocker monotherapy, and calcium channel blocker/ angiotensin converting enzyme inhibitor or angiotensin receptor blocker dual therapy.

Approximately seventy percent of the subjects admitted for acute stroke had a documented history of hypertension. In contrast, only half of the subjects were taking blood pressure agents prior to admission because many patients had stopped taking their medications. Underlying reasons for discontinuing antihypertensives could not be ascertained due to limited documentation, but unaffordability and loss to outpatient follow up likely contributed to treatment discontinuation. For the patients taking antihypertensive medications, the selection of drug combinations by providers was consistent with the ALLHAT trial’s recommendation for calcium channel blockers, thiazide diuretics and ACE or ARBs as first-line blood pressure agents.^*12*^

A higher mortality of 67% (n=9) compared to 40% (n=10) among males and females taking three or more outpatient hypertensive medications respectively. This difference was likely due to the small sizes of these subgroups.

The similarity in mortality outcomes between patients taking antihypertensives versus patients not taking blood pressure medications prior to acute stroke onset is likely multifactorial. First, it is likely that many of the patients taking antihypertensives had poorly controlled hypertension. A study on hypertension management in Zimbabwe showed a 78% blood pressure medication adherence with only 22% optimal blood pressure control.^***13***^ Second, a relatively high number of patients presenting with hemorrhagic stroke in both groups could have contributed to similarities in mortality outcomes due to the high upfront acute mortality rate of hemorrhagic strokes. Worldwide, about 10% of strokes present with hemorrhage and carry a 4-fold increase in 7-day mortality compared to ischemic strokes.^***14***^ A 100-patient case-study on computed tomography imaging findings in Zimbabwean stroke patients showed that 29% of stroke presentations were hemorrhagic at a single center.^***11***^ Despite the case-study’s small sample size, it may be inferred that Zimbabweans may be at a comparatively higher risk for presenting with hemorrhagic stroke. The inherent mortality risk from hemorrhagic stroke could have superseded any benefits from prior outpatient blood pressure management. The number of hemorrhagic strokes in this study could not be obtained due to lack of computed tomography or magnetic resonance imaging at our center.

### Comparison to the 2017 Harare Stroke Study

Historically, most of the stroke studies in Zimbabwe have been performed in the better-resourced and more densely populated capital city, Harare. Therefore, an additional objective of this study was to compare our data to the recent 2017 stroke study that was performed in three Harare teaching hospitals.^***8***^ Two main similarities in outcomes for both studies were the female predominance in stroke patients and the mean age of stroke presentation in the 60s. In contrast, our study showed a higher prevalence of prior hypertension and higher inpatient mortality from acute stroke compared to the observations in the 2017 Harare study.

Stroke disproportionately affects females more than males across Zimbabwe. Females accounted for about two thirds of the subjects in both studies. This over-representation among acute stroke patients does not reflect the national population distribution by sex in which females account for 51.9% of the general population.^***15***^ Mean age of stroke presentation was consistent across both studies. Average of subjects in our study was 65.8 ± 15.7 years in our study compared to 61.6 ± 16.8 years in the Harare study.

There was a notable 10.6% difference in the prevalence of hypertension noted across both studies. In our investigation, the hypertension prevalence was higher at 69.0% compared to 58.4% in the Harare study.

Overall inpatient mortality in our study was 35.5% which was notably higher than the 24.9% inpatient mortality reported in the multi-center Harare study. Of the three centers included in the Harare study, Parirenyatwa is most similar to our center in terms of patient volume and acuity of medical care. Inpatient stroke mortality at Parirenyatwa was 26.3% compared to 35.5% in our study. Several disparities in acute stroke care exist between the two centers. Compared to our center, Parirenyatwa has a dedicated stroke care unit, access to computed tomography neuroimaging, and a neurosurgery service that is available for emergent decompression of hemorrhagic strokes with sequelae of increased intracranial pressure or acute obstructive hydrocephalus.

### Limitations

This study had several limitations. First, inadequate and non-uniform documentation limited the gathering of sufficient accurate data. A total of 28 subjects had to be excluded for inadequately charted information. Lack of standard history and physical templates limits the ability to consistently collect data for research. Second, the inability to access outpatient medical records limited our ability to determine the proportion of patients on optimal blood pressure control prior to acute stroke onset. Additionally, one third of patients taking antihypertensive medications did not recall the names of the medications they were taking at home. Universal knowledge of the home blood pressure regimens would have provided a more accurate account of the most common hypertension regimens for patients admitted for stroke in this study. Lastly, the lack of neuro-imaging and electro-encephalogram data limited our ability to definitively rule out technically challenging stroke mimics such as intracranial tumors, brain abscesses, and non-convulsive status epilepticus.

## Conclusion

Approximately half of all patients admitted for stroke were taking antihypertensive medications despite a 69.0% prevalence of hypertension. There was no statistical difference in mortality among patients taking or not taking antihypertensives prior to stroke onset. The most common antihypertensive regimens were CCB/thiazide dual therapy, thiazide monotherapy, CCB monotherapy, and CCB/ACE-inhibitors or ARB dual therapy respectively. Despite similar demographics, disparities in distribution of resources for acute stroke exist across tertiary centers in Zimbabwe. The data obtained from this study was pivotal in advocating for improved acute stroke care in Southern Zimbabwe and led to the opening of a new stroke ward at our center in 2021. A standardized template for admissions to this new unit has been generated to facilitate uniform documentation.

Future research is required to evaluate inpatient mortality outcomes among patients with well-controlled versus uncontrolled hypertension prior to acute stroke. Additionally, a follow up study on acute stroke inpatient mortality from the new stroke ward will provide a pre-and post-intervention analysis when juxtaposed with the current data.

## Data Availability

All raw data is available via email in Excel spreadsheet format upon reasonable request.

